# Age-related changes in the upper respiratory microbiome are associated with SARS-CoV-2 susceptibility and illness severity

**DOI:** 10.1101/2021.03.20.21252680

**Authors:** Jillian H. Hurst, Alexander W. McCumber, Jhoanna N. Aquino, Javier Rodriguez, Sarah M. Heston, Debra J. Lugo, Alexandre T. Rotta, Nicholas A. Turner, Trevor S. Pfeiffer, Thaddeus C. Gurley, M. Anthony Moody, Thomas N. Denny, John F. Rawls, Christopher W. Woods, Matthew S. Kelly

## Abstract

Children are less susceptible to SARS-CoV-2 and typically have milder illness courses than adults. We studied the nasopharyngeal microbiomes of 274 children, adolescents, and young adults with SARS-CoV-2 exposure using 16S rRNA gene sequencing. We find that higher abundances of *Corynebacterium* species are associated with SARS-CoV-2 infection and SARS-CoV-2-associated respiratory symptoms, while higher abundances of *Dolosigranulum pigrum* are present in SARS-CoV-2-infected individuals *without* respiratory symptoms. We also demonstrate that the abundances of these bacteria are strongly, and independently, associated with age, suggesting that the nasopharyngeal microbiome may be a potentially modifiable mechanism by which age influences SARS-CoV-2 susceptibility and severity.

**Summary:** Evaluation of nasopharyngeal microbiome profiles in children, adolescents, and young adults with a SARS-CoV-2-infected close contact identified specific bacterial species that vary in abundance with age and are associated with SARS-CoV-2 susceptibility and the presence of SARS-CoV-2-associated respiratory symptoms.

---

SARS-CoV-2, the etiological agent of coronavirus disease 2019 (COVID-19), has resulted in more than 100 million infections and two million deaths globally [1]. In contrast to most other respiratory viruses [2-4], children appear to be less susceptible to SARS-CoV-2 infection, and SARS-CoV-2-infected children typically have milder illness courses than adults. In a recent meta-analysis that included 32 studies of 41,640 children and adolescents less than 20 years of age and 268,945 adults 20 years of age or older, children and adolescents were estimated to have a 46% lower susceptibility to SARS-CoV-2 infection than adults [5]. Further, a higher incidence of SARS-CoV-2 infection has been observed with increasing age, even among infants, children, and adolescents [6]. Reporting data from a community-based study of SARS-CoV-2 infections, we previously demonstrated that at least one-third of SARS-CoV-2-infected children and adolescents are asymptomatic [7], and children who do develop symptomatic SARS-CoV-2 infection typically report mild respiratory symptoms [7-10]. Additionally, COVID-19 hospitalization rates and mortality among children are substantially lower than among adults of all ages, with only 576 hospitalizations and 208 deaths among children and adolescents less than 18 years of age reported during the first six months of the pandemic in the United States [11]. These data suggest that host biological or immunological factors that vary with age modify susceptibility to and severity of SARS-CoV-2 infection, and that an understanding of these host factors could inform development of novel strategies to prevent or treat SARS-CoV-2 infections in children and adults.

Given that the upper respiratory microbiome undergoes substantial shifts in early childhood [12-14], and is increasingly recognized to play a key role in the pathogenesis of respiratory virus infections [15-21], we hypothesized that changes in the upper respiratory microbiome with age might contribute to the differences in susceptibility to SARS-CoV-2 infection and the severity of SARS-CoV-2-associated respiratory symptoms among children and adults. In this study, we used 16S ribosomal RNA (rRNA) gene amplicon sequencing to characterize the nasopharyngeal microbiomes of 274 children, adolescents, and young adults with close contact with a SARS-CoV-2-infected individual. We describe associations between the composition of the nasopharyngeal microbiome and SARS-CoV-2 infection. In addition, we identify bacterial species that are associated with the presence of respiratory symptoms among children with SARS-CoV-2 infection. Finally, we demonstrate that the relative abundances of these bacterial species within the nasopharyngeal microbiome are strongly, and independently, associated with age. Taken together, our findings demonstrate that age-associated changes in the upper respiratory microbiome may contribute to the varied susceptibility to and severity of SARS-CoV-2 infection among children and adults.

## METHODS

### Study Population

The Duke Biospecimens from RespirAtory Virus-Exposed Kids (BRAVE Kids) study is a prospective cohort study of children, adolescents, and young adults with confirmed SARS-CoV-2 infection or close contact with an individual with confirmed SARS-CoV-2 infection [7]. This study is being conducted within the Duke University Health System in Raleigh-Durham, North Carolina. Eligible participants are less than 21 years of age and have close contact with an individual with SARS-CoV-2 infection confirmed by real-time PCR. Close contact is defined as an unprotected exposure within 6 feet to a confirmed case between 2 days before and 7 days after symptom onset or confirmation of SARS-CoV-2 infection in asymptomatic contacts. Close contacts include, but are not limited to, parents, siblings, other caregivers, and partners.

### Study Procedures

We collected exposure, sociodemographic, and clinical data at enrollment through review of electronic medical records and a directed questionnaire conducted by telephone. We recorded any symptoms reported within the 2 weeks preceding study enrollment. In addition, we conducted follow-up questionnaires for all participants 7 days after study enrollment to document new symptoms and healthcare encounters. For participants with one or more symptoms 7 days after study enrollment, additional questionnaires were administered 14 and 28 days after enrollment, or until the participant reported complete symptom resolution. We recorded the results of SARS-CoV-2 testing performed for clinical care. In addition, we conducted a home visit as soon as possible after study enrollment to collect a nasopharyngeal swab and other biospecimens. Participants who declined a home visit received a kit for self-collection of a mid-turbinate nasal swab. Nasopharyngeal and nasal samples were collected with nylon flocked swabs (Copan Italia, Brescia, Italy) and placed into RNAProtect (Qiagen, Hilden, Germany) prior to storage at −80°C. All nasopharyngeal and nasal samples were tested for SARS-CoV-2 using a real-time quantitative PCR assay, as previously described [7]. We classified participants as SARS-CoV-2-infected if SARS-CoV-2 was identified in either a clinical or research PCR assay. We considered SARS-CoV-2-infected individuals as having respiratory symptoms if they reported cough, rhinorrhea, nasal congestion, shortness of breath, sore throat, or anosmia at any point between 14 days prior to enrollment through the end of the study follow-up period. Finally, we classified participants into one of three groups: 1) SARS-CoV-2-uninfected; 2) SARS-CoV-2-infected *without* respiratory symptoms; and 3) SARS-CoV-2-infected *with* respiratory symptoms. The analyses presented herein were limited to study participants with an available nasopharyngeal sample.

### Processing of nasopharyngeal samples for 16S ribosomal RNA sequencing

The Duke Microbiome Core Facility extracted DNA from nasopharyngeal samples using Powersoil Pro extraction kits (Qiagen) following the manufacturer’s instructions. DNA concentrations were determined using Qubit dsDNA high-sensitivity assay kits (Thermo Fisher Scientific). Bacterial community composition was characterized by PCR amplification of the V4 variable region of the 16S rRNA gene using the forward primer 515 and the reverse primer 806 following the Earth Microbiome Project protocol [22]. These primers carry unique barcodes that allow for multiplexed sequencing. Equimolar 16S rRNA PCR products from all samples were quantified and pooled prior to sequencing. Sequencing was performed by the Duke Sequencing and Genomic Technologies Core Facility on an Illumina MiSeq instrument configured for 250 base-pair paired-end sequencing. All samples were included in a single sample processing run with inclusion of both negative extraction and PCR controls. We analyzed raw sequences through a DADA2 pipeline [23] and assigned taxonomy to amplicon sequence variants (ASVs) using Silva SSU version 138 [24]. We identified and removed sequencing contaminants (n=40) using the *decontam* R package [25]. Samples with less than 1000 sequencing reads after quality filtering and contaminant removal were excluded. We obtained a median [interquartile range (IQR)] of 24,422 (17,799, 33,291) high-quality sequencing reads from the 274 samples included in these analyses. Sequencing reads were classified into 1,799 ASVs representing 316 bacterial genera from 20 phyla (**Supplemental Table 1**). We performed standard nucleotide REFSEQ BLAST searches of ASV reference sequences using the National Center for Biotechnology Information’s Bacteria and Archaea 16S ribosomal RNA project database [26]. We assigned species information to ASVs using a best-hit approach based on the E value with a minimum percent identity of 99%.

### Data Analysis

We first sought to characterize changes in nasopharyngeal microbiome diversity and composition that occur with age. We calculated nasopharyngeal microbiome alpha (Shannon and Chao1 indices) and beta diversity (Bray-Curtis dissimilarity) using the *phyloseq* R package [27]. We then fit linear regression models to evaluate associations between participant age and microbiome alpha diversity measures. The Chao1 index was not normally distributed and was thus log-transformed for these analyses. We then evaluated the association between age and nasopharyngeal microbiome composition with PERMANOVA using the adonis function within the *vegan* R package [28]. Finally, we used ANCOM-II [29] to fit linear models evaluating associations between age and the relative abundances of bacterial ASVs within the nasopharyngeal microbiome. An ASV was considered to be differentially abundant if it varied across the independent variable of interest with respect to 80% of the other ASVs in the dataset (*W* statistic > 0.80). ASVs that were identified as structural zeros were not considered to be differentially abundant. All analyses of nasopharyngeal microbiome diversity and composition by age were adjusted for participant SARS-CoV-2 infection status and the presence of SARS-CoV-2-associated respiratory symptoms. We then used these same methods to evaluate associations between nasopharyngeal microbiome diversity and composition and SARS-CoV-2 infection status, adjusting for participant age. Finally, in analyses limited to SARS-CoV-2-infected individuals, we used these same methods to evaluate associations between the nasopharyngeal microbiome and the presence of respiratory symptoms among SARS-CoV-2-infected participants, adjusting for age. Our findings were not substantively changed when we additionally adjusted for sex and race in analyses of nasopharyngeal microbiome diversity and composition by age, SARS-CoV-2 status, or SARS-CoV-2-associated respiratory symptoms. Given that ANCOM-II produces only a list of differentially abundant ASVs, we used DESeq2 [30] to generate log2-fold changes in abundance in order to determine the directionality of these associations. Analyses conducted in ANCOM-II and DESeq2 were limited to ASVs present in at least 5% of samples and adjusted for the false discovery rate using the Benjamini-Hochberg procedure. Analyses were performed using R version 4.0.3 [31] and all visualizations were created using the *ggplot2* R package [32].

## RESULTS

### Characteristics of the study population

Two hundred seventy-four children, adolescents, and young adults were included in these analyses (**Table 1**). Participants were classified as SARS-CoV-2-uninfected (n=75, 27%); SARS-CoV-2-infected *without* respiratory symptoms (n=88, 32%); and SARS-CoV-2-infected *with* respiratory symptoms (n=111, 41%). Among SARS-CoV-2-infected individuals, nasopharyngeal samples were collected a median (IQR) of 5 (2, 8) days from symptom onset (or SARS-CoV-2 diagnosis in asymptomatic cases). SARS-CoV-2-infected participants *with* respiratory symptoms were older than SARS-CoV-2-infected participants without respiratory symptoms and SARS-CoV-2-uninfected participants, consistent with our previous observation of a high prevalence of respiratory symptoms among SARS-CoV-2-infected adolescents in this cohort [7]. Similarly, a higher proportion of SARS-CoV-2-infected individuals identified as Latino or Hispanic-American compared to SARS-CoV-2-uninfected individuals [7]. There were no significant differences in the prevalences of comorbidities or recent receipt of antibiotics or probiotics in these groups.

**Table 1.**
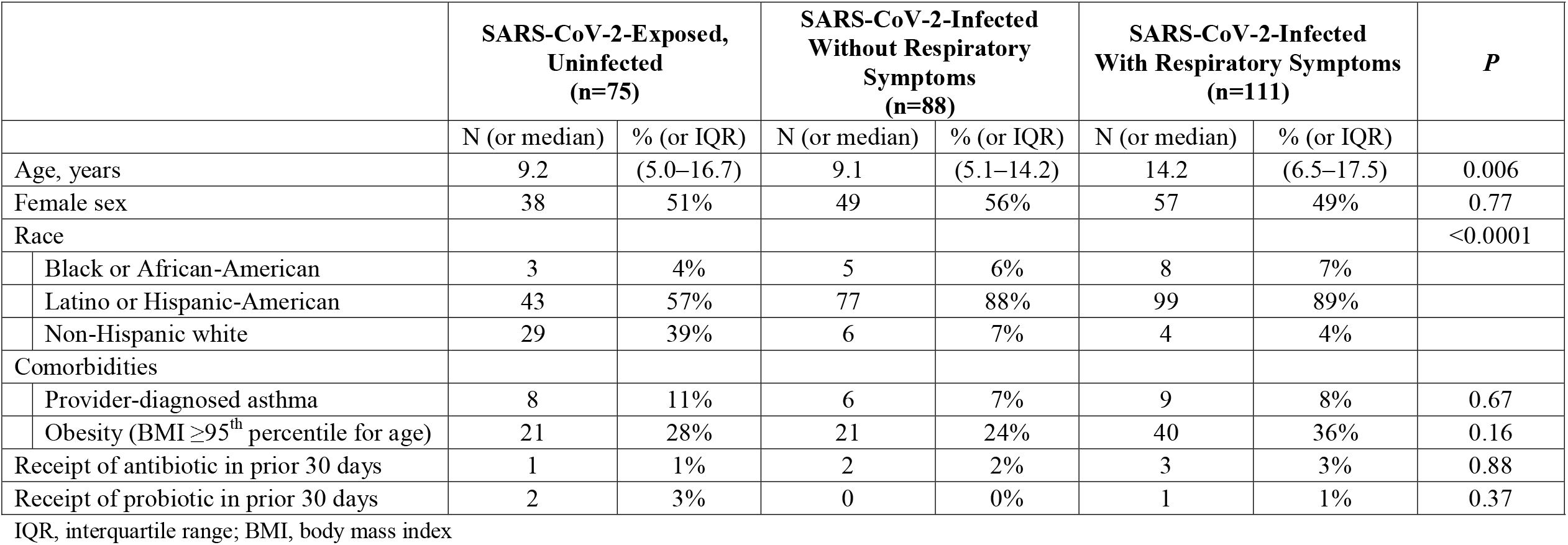
Characteristics of the study population

|  | **SARS-CoV-2-Exposed,**<br>**Uninfected**<br>**(n=75)** |  | **SARS-CoV-2-Infected**<br>**Without Respiratory**<br>**Symptoms**<br>**(n=88)** |  | **SARS-CoV-2-Infected**<br>**With Respiratory Symptoms**<br>**(n=111)** |  | ***P*** |
| --- | --- | --- | --- | --- | --- | --- | --- |
|  | N (or median) | % (or IQR) | N (or median) | % (or IQR) | N (or median) | % (or IQR) |  |
| Age, years | 9.2 | (5.0–16.7) | 9.1 | (5.1–14.2) | 14.2 | (6.5–17.5) | 0.006 |
| Female sex | 38 | 51% | 49 | 56% | 57 | 49% | 0.77 |
| Race |  |  |  |  |  |  | <0.0001 |
| Black or African-American | 3 | 4% | 5 | 6% | 8 | 7% |  |
| Latino or Hispanic-American | 43 | 57% | 77 | 88% | 99 | 89% |  |
| Non-Hispanic white | 29 | 39% | 6 | 7% | 4 | 4% |  |
| Comorbidities |  |  |  |  |  |  |  |
| Provider-diagnosed asthma | 8 | 11% | 6 | 7% | 9 | 8% | 0.67 |
| Obesity (BMI ≥95<sup>th</sup> percentile for age) | 21 | 28% | 21 | 24% | 40 | 36% | 0.16 |
| Receipt of antibiotic in prior 30 days | 1 | 1% | 2 | 2% | 3 | 3% | 0.88 |
| Receipt of probiotic in prior 30 days | 2 | 3% | 0 | 0% | 1 | 1% | 0.37 |

IQR, interquartile range; BMI, body mass index

### Age-associated changes in nasopharyngeal microbiome composition

Median (IQR) Shannon diversity and Chao1 indices of the nasopharyngeal microbiome were 1.49 (1.12, 2.00) and 70.0 (48.8, 97.2), respectively. Nasopharyngeal microbiome diversity, as measured by the Shannon diversity index, increased with age (**Figure 1a**; p<0.0001), while nasopharyngeal microbiome richness, as measured by the Chao1 index, declined with increasing age (**Figure 1b**; p=0.02). The composition of the nasopharyngeal microbiome also differed according to age (**Figure 1c**; PERMANOVA on Bray-Curtis dissimilarity, p<0.001, R^2^=0.103). In particular, older age was associated with lower abundances of several ASVs classified as *Dolosigranulum, Moraxella*, and *Streptococcus*, and higher abundances of specific ASVs classified as *Anaerococcus, Corynebacterium, Lawsonella, Peptoniphilus*, and *Staphylococcus* (**Figure 2** and **Supplemental Table 2**). Infants and young children 5 years of age or younger tended to have high abundances of *Dolosigranulum* and *Moraxella* within their nasopharyngeal microbiomes, while the nasopharyngeal microbiomes of adolescents and young adults 12 years of age or older were frequently dominated by *Staphylococcus*. Additionally, several bacterial genera (*Anaerococcus, Lawsonella*, and *Peptoniphilus*) that were of low prevalence and abundance in the nasopharyngeal microbiomes of infants and young children were highly abundant in adolescents and young adults (**Supplemental Table 3**). *Corynebacterium* was highly abundant in the nasopharyngeal microbiome across all age categories and its relative abundance was positively correlated with age.

**Figure 1.**
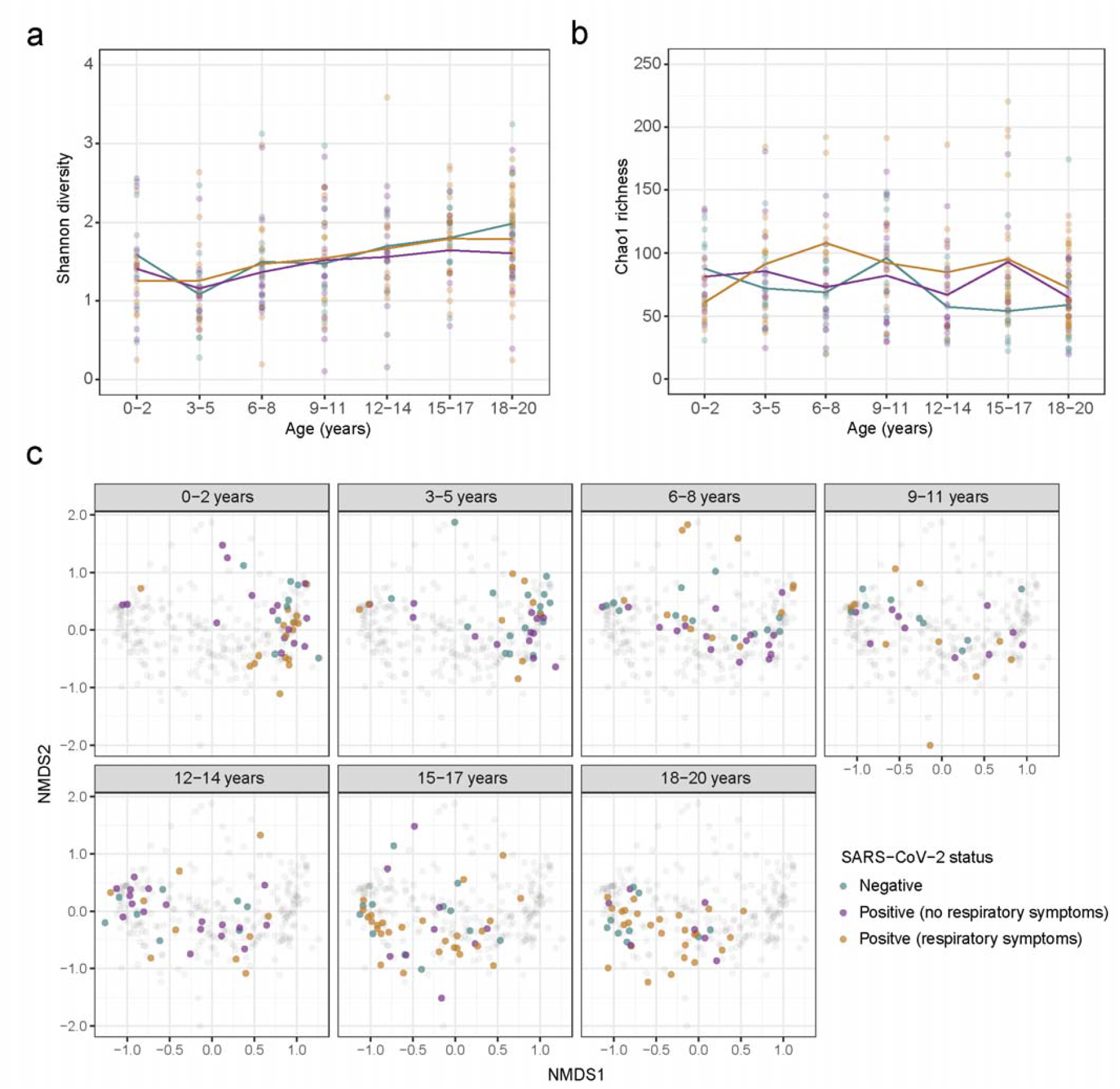
Nasopharyngeal microbiome alpha and beta diversity by age and SARS-CoV-2 clinical status. Shannon diversity (a) and Chao1 richness (b) is shown for each age group by clinical status: SARS-CoV-2-uninfected (green), SARS-CoV-2-infected *without* respiratory symptoms (purple), and SARS-CoV-2-infected *with* respiratory symptoms (orange). Points represent individual samples while the line denotes means for samples from each group in each age category. (c) Non-metric multidimensional scaling plot based on Bray-Curtis distances is shown by age category and according to SARS-CoV-2 clinical status [(SARS-CoV-2-uninfected (green), SARS-CoV-2-infected *without* respiratory symptoms (purple), and SARS-CoV-2-infected *with* respiratory symptoms (orange)]. Within each panel corresponding to an age category, the colored points represent samples for the corresponding age category, while the gray points represent samples collected from children in other age categories.

**Figure 2.**
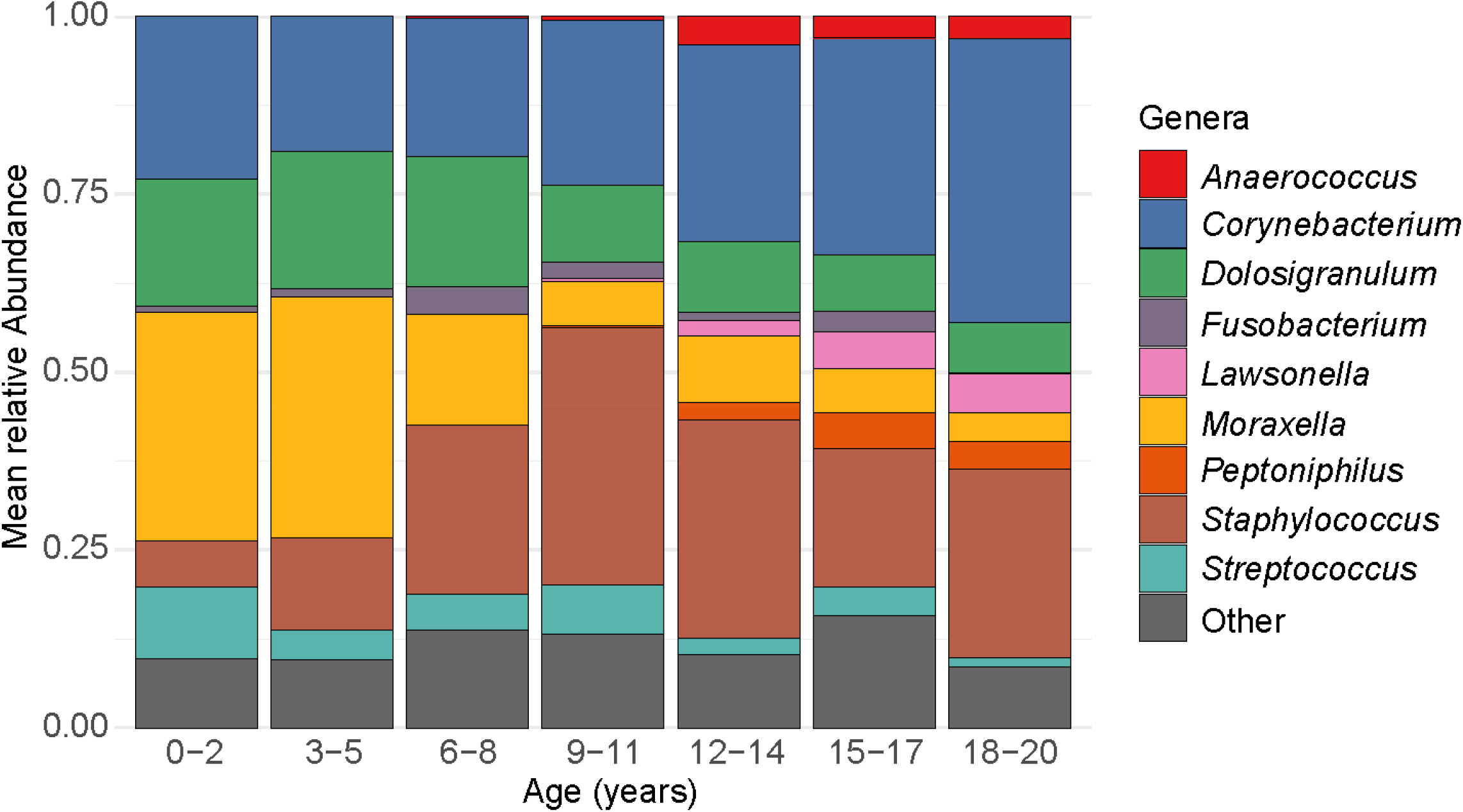
Relative abundances of highly abundant genera by age. Each bar depicts the mean relative abundances of highly abundant genera in nasopharyngeal samples from participants in a specific age category. Only the nine most highly abundant genera within nasopharyngeal samples from the entire study population are shown.

### Associations between the nasopharyngeal microbiome and SARS-CoV-2 susceptibility

Nasopharyngeal microbiome diversity, as measured by the Shannon diversity index, did not differ in SARS-CoV-2-infected and SARS-CoV-2-uninfected participants (**Figure 1a**; p=0.36). In contrast, SARS-CoV-2-infected individuals had higher microbiome richness, measured by the log-transformed Chao1 index, than uninfected individuals (**Figure 1b**; p=0.008). Additionally, nasopharyngeal microbiome composition differed in SARS-CoV-2-infected and uninfected participants (**Figure 1c**; PERMANOVA on Bray-Curtis dissimilarity, p=0.007, R^2^=0.009). In ANCOM-II analyses adjusting for age, we identified two ASVs assigned to the bacterial genus *Corynebacterium* that were differentially abundant by SARS-CoV-2 infection status (**Figure 3a** and **Supplemental Table 4**), both of which were of higher abundance in SARS-CoV-2-infected participants relative to uninfected participants. Using BLAST searches, we operationally classified these ASVs as *Corynebacterium accolens/macginleyi/tuberculostearicum*. Notably, the relative abundances of both of these ASVs increased with increasing age independent of SARS-CoV-2 infection status (**Figure 3b** and **Supplemental Table 2**).

**Figure 3.**
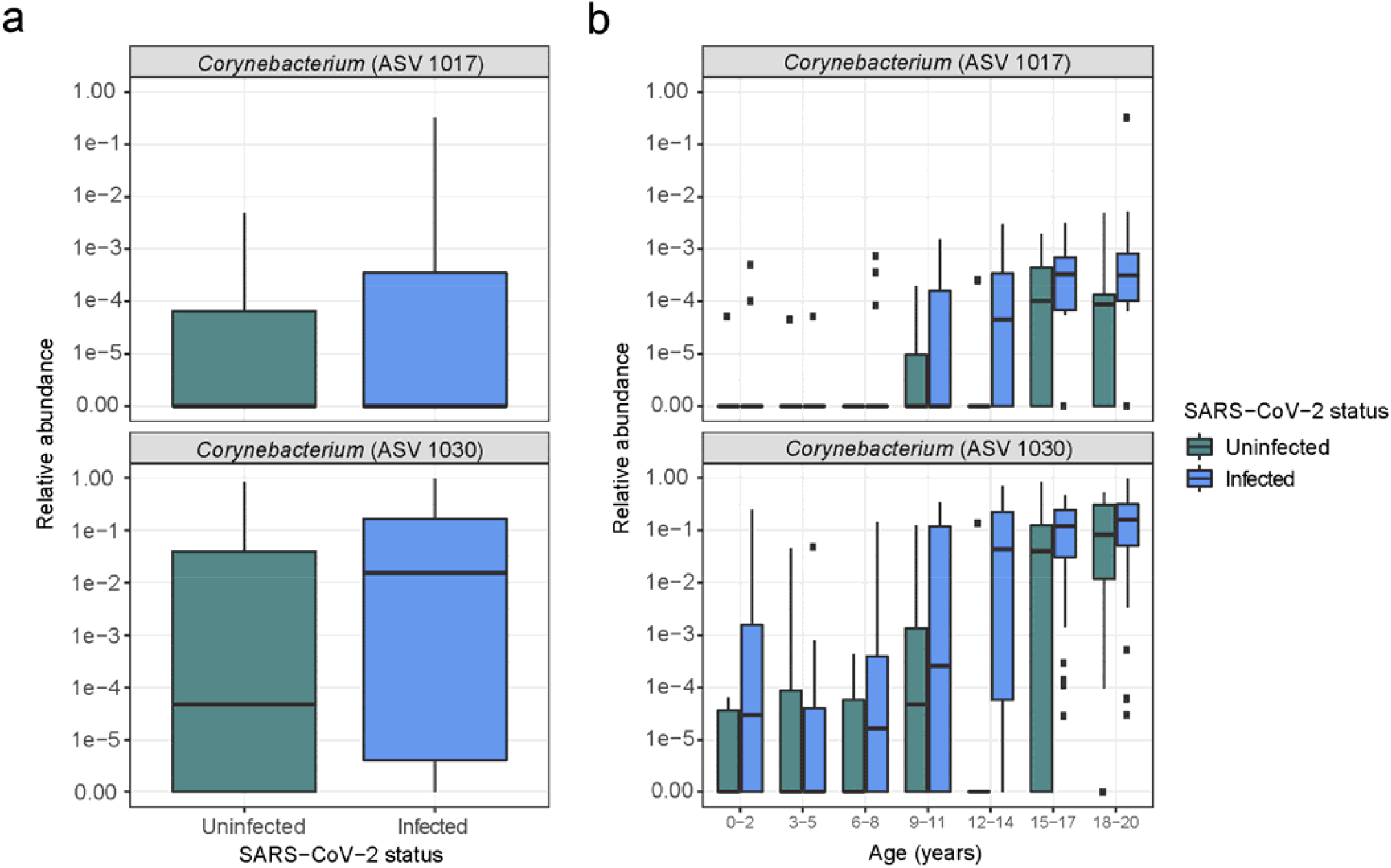
Relative abundances of ASVs that were differentially abundant by SARS-CoV-2 infection status. a) Boxplots of the relative abundances of ASVs that were differentially abundant by SARS-CoV-2 infection status in age-adjusted analyses conducted in ANCOM-II. a) Boxplots of the relative abundances by age group of ASVs that were differentially abundant by SARS-CoV-2 infection status in age-adjusted analyses conducted in ANCOM-II. Relative abundances are plotted on a log scale with 1E-6 added to all counts to enable transformation of zero counts. Rectangles represent the lower quartile, median, and upper quartile of the relative abundances of each ASV. The relative abundances of two ASVs classified as *Corynebacterium accolens/macginleyi/tuberculostearicum* were higher in SARS-CoV-2-infected individuals compared to uninfected individuals, adjusting for age (ASV 1017, *W*=192; ASV 1030, *W*=233). In addition, the relative abundances of these *Corynebacterium* ASVs increased with increasing age adjusting for SARS-CoV-2 infection status and the presence of SARS-CoV-2-associated respiratory symptoms (ASV 1017, *W*=170; ASV 1030, *W*=173).

### Associations between the nasopharyngeal microbiome and SARS-CoV-2-associated respiratory symptoms

SARS-CoV-2 infection results in a broad range of clinical manifestations from completely asymptomatic to severe pneumonia and acute respiratory distress syndrome [33-35]. In particular, we previously reported marked differences in the prevalences of respiratory symptoms among children, adolescents, and young adults with SARS-CoV-2 infection [7]. To evaluate the extent to which the upper respiratory microbiome contributes to the varied clinical presentations of SARS-CoV-2 infection by age, we compared the nasopharyngeal microbiomes of SARS-CoV-2-infected participants *with* respiratory symptoms to those of SARS-CoV-2-infected participants *without* respiratory symptoms. Nasopharyngeal microbiome alpha diversity (Shannon diversity, Chao1 richness) was similar in SARS-CoV-2-infected individuals with or without respiratory symptoms (**Figure 1a** and **Figure 1b**), while nasopharyngeal microbiome composition differed between these groups (**Figure 1c**; PERMANOVA on Bray-Curtis dissimilarity, p=0.008, R^2^=0.014). In ANCOM-II analyses adjusting for age, we identified nine ASVs that were differentially abundant in SARS-CoV-2-infected participants with or without respiratory symptoms (**Figure 4a** and **Supplemental Table 5**). The two ASVs classified as *Corynebacterium* that were associated with SARS-CoV-2 infection status (ASV 1017, ASV 1030) were also more abundant in SARS-CoV-2-infected individuals *with* respiratory symptoms compared to SARS-CoV-2-infected individuals *without* respiratory symptoms. Additionally, two other *Corynebacterium* ASVs that were identified as *Corynebacterium accolens/macginleyi/tuberculostearicum* (ASV 1014, ASV 1032) were more abundant in SARS-CoV-2-infected participants *with* respiratory symptoms. Other ASVs that were more abundant in SARS-CoV-2-infected participants *with* respiratory symptoms were classified as *Lawsonella clevelandensis* (ASV 1060), *Finegoldia magna* (ASV 1535), *Anaerococcus* spp. (ASV 1593) and *Peptononiphilus* spp. (ASV 1618). Each of the ASVs that were associated with the presence of respiratory symptoms in SARS-CoV-2-infected individuals were independently and positively associated with increasing age (**Figure 4b** and **Supplemental Table 2**), with several ASVs that were almost exclusively detected in adolescents 12 years of age or older. We also identified a single ASV that was assigned to *Dolosigranulum pigrum* (ASV 1491) that was of lower abundance in SARS-CoV-2-infected participants with respiratory symptoms and additionally declined in abundance with increasing age. Finally, nasopharyngeal microbiome composition also differed between SARS-CoV-2-infected participants *with* respiratory symptoms and SARS-CoV-2-uninfected participants (PERMANOVA on Bray-Curtis dissimilarity, p<0.001, R^2^=0.015), with several of these same *Corynebacterium* ASVs (ASV 1017, ASV 1030, ASV 1032) and ASVs classified as *Anaerococcus* spp. (ASV 1582) and *Peptoniphilus* spp. (ASV 1618) being more abundant in the nasopharyngeal microbiomes of SARS-CoV-2-infected participants *with* respiratory symptoms (**Supplemental Table 6**).

**Figure 4.**
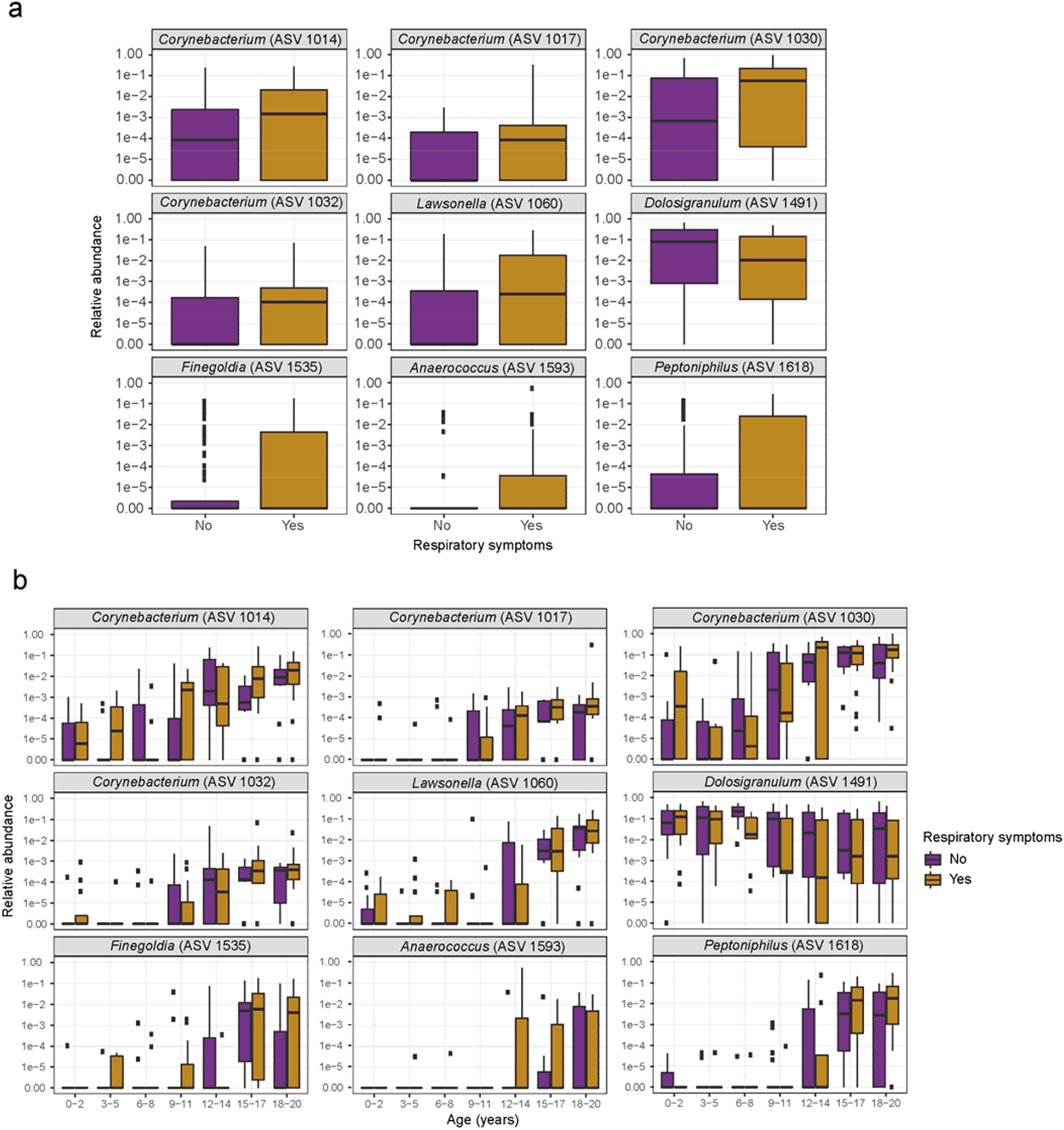
Relative abundances of ASVs that were differentially abundant by SARS-CoV-2-associated respiratory symptoms. a) Boxplots of the relative abundances of ASVs that were differentially abundant by the presence of respiratory symptoms in SARS-CoV-2-infected individuals in age-adjusted analyses performed in ANCOM-II. b) Boxplots of the relative abundances by age group of ASVs that were differentially abundant by SARS-CoV-2 infection status in age-adjusted analyses conducted in ANCOM-II. Relative abundances are plotted on a log scale with 1E-6 added to all counts to enable transformation of zero counts. Rectangles represent the lower quartile, median, and upper quartile of the relative abundances of each ASV. Adjusting for age, SARS-CoV-2-associated respiratory symptoms were associated with higher abundances of ASVs assigned to *Corynebacterium* spp. (ASV 1014, *W*=268; ASV 1017, *W*=233; ASV 1030, *W*=271; ASV 1032, *W*=229), *Lawsonella clevelandensis* (ASV 1060, *W*=273), *Finegoldia magna* (ASV 1535, *W*=265), *Anaerococcus* spp. (ASV 1593, *W*=231), and *Peptoniphilus* spp. (ASV 1618, *W*=271). The relative abundances of each of these ASVs additionally increased with increasing age adjusting for SARS-CoV-2 infection status and the presence of SARS-CoV-2-associated respiratory symptoms (ASV 1014, *W*=173; ASV 1017, *W*=170; ASV 1030, *W*=173; ASV 1032, *W*=170; ASV 1060, *W*=173; ASV 1535, *W*=172; ASV 1593, *W*=169; ASV 1618, *W*=173). Adjusting for age, SARS-CoV-2-associated respiratory symptoms were associated with a lower abundance of a single ASV assigned to *Dolosigranulum pigrum* (ASV 1491, *W*=260), and the relative abundance of this ASV decreased with increasing age adjusting for SARS-CoV-2 infection status and the presence of SARS-CoV-2-associated respiratory symptoms (ASV 1491, *W*=171).

## DISCUSSION

In this study, we found that higher abundances of specific *Corynebacterium* ASVs are associated with SARS-CoV-2 infection among children, adolescents, and young adults with a SARS-CoV-2-infected close contact. In addition, we identified nasopharyngeal microbiome alterations, including higher abundances of *Corynebacterium* and lower abundances of *D. pigrum*, that are associated with the presence of respiratory symptoms in the setting of confirmed SARS-CoV-2 infection. Finally, we demonstrate that these same microbiome features are strongly, and independently, associated with increasing age. Our findings suggest that the upper respiratory microbiome may be a previously unrecognized and potentially modifiable mechanism by which age influences SARS-CoV-2 susceptibility and illness severity.

Over the past several decades, accumulating data have emerged demonstrating that the upper respiratory microbiome plays a key role in the pathogenesis of respiratory virus infections. First, several studies indicate that the upper respiratory microbiome modifies susceptibility to respiratory virus infections, as has been most clearly demonstrated for influenza viruses through household transmission studies [15, 16]. Among 436 household contacts in one of these studies, higher abundances of *Streptococcus* spp. and *Prevotella salivae* within the nasal/oropharyngeal microbiome were associated with lower susceptibility to influenza A infection [15]. Additionally, the upper respiratory microbiome may also influence the symptoms experienced by individuals when respiratory virus infection does occur, presumably through modulation of the host immune response [36]. For instance, the nasopharyngeal microbiome appears to modify the severity of respiratory syncytial virus (RSV) infection among young children, with several studies reporting that higher abundances of *Haemophilus* species are associated with a more exuberant host immune response to RSV [17-19]. Nasopharyngeal microbiome profiles are also associated with the levels of inflammatory cytokines in nasal wash samples and the development of symptomatic infection in adults following experimental rhinovirus challenge [20, 21]. Additionally, intranasal administration of live bacterial strains has been shown to directly modulate immune responses to influenza viruses and RSV in animal models [37-40]. While data from clinical studies are currently lacking, this work illustrates the potential that targeted manipulation of the upper respiratory microbiome holds for the prevention and treatment of respiratory virus infections.

To date, studies of the upper respiratory microbiome and SARS-CoV-2 infection have been conducted among small cohorts of adults presenting with clinical suspicion of COVID-19. De Maio and colleagues did not find any differences in the nasopharyngeal microbiomes of 18 individuals with symptomatic SARS-CoV-2 infection and 22 individuals with acute respiratory symptoms who tested negative for SARS-CoV-2 [41]. Conversely, a study of 59 adults identified several ASVs that were differentially abundant in those with SARS-CoV-2 infection and in those with higher viral loads during COVID-19, including higher abundances of ASVs classified as *Peptoniphilus lacrimalis, Prevotella copri*, and *Anaerococcus* spp. [42]. By comparison, Mostafa and colleagues reported lower nasopharyngeal microbial diversity, a lower abundance of the bacterial family Propionibacteriaceae, and a higher abundance of *Corynebacterium accolens* in 40 SARS-CoV-2-infected adults compared to 10 SARS-CoV-2-uninfected adults with suspected COVID-19 [43]. Consistent with findings from this latter study, and in the largest cohort studied to date, here we demonstrated that ASVs assigned to *Corynebacterium* spp. (*C. accolens/macginleyi/tuberculostearicum*) were associated with SARS-CoV-2 infection among 274 children, adolescents, and adults with SARS-CoV-2 exposure. In addition, we identified novel associations between ASVs assigned to *Corynebacterium* spp., *Lawsonella clevelandensis, Finegoldia magna, Anaerococcus* spp., *Peptoniphilus* spp., and *Dolosigranulum pigrum* and the presence of respiratory symptoms during SARS-CoV-2 infection. Notably, the changes in relative abundances of these ASVs that were associated with SARS-CoV-2 infection and SARS-CoV-2-associated respiratory symptoms were also independently associated with increasing age, suggesting that age-associated changes in the upper respiratory microbiome may underlie the differences in SARS-CoV-2 susceptibility and illness severity that exist between children and adults.

Non-*diphtheriae Corynebacterium* species and *D. pigrum* were previously shown to have important microbial interactions within the human nasopharynx. In particular, the abundance of *Corynebacterium* within the nasopharyngeal microbiome has been negatively associated with colonization by *Streptococcus pneumoniae* among infants and children [44-47], while the relative abundance of *D. pigrum* within the nasopharyngeal microbiome has been negatively associated with the co-occurrence of both *S. pneumoniae* and *S. aureus* [47, 48]. Few prior clinical studies identified associations between these bacterial species and the risk and severity of respiratory virus infections. Interestingly, strains of *Corynebacterium* spp. and *D. pigrum* have been shown to influence innate immune responses to influenza and RSV in murine models [49-51], suggesting that these bacterial commensals may influence overall immunological tone within the upper respiratory tract. Importantly, much of the work on these bacterial species has been conducted in young children [44-48], and surprisingly little is known about the abundances and functions of these microbes in the upper respiratory tracts of adults. Taken together, these studies suggest that specific strains of commensal bacteria have differential impacts on host immune responses to potential pathogens and that age-associated changes in the relative abundance of different species likely plays a critical role in infection susceptibility.

Previous studies demonstrated that the nasopharyngeal microbiome is a relatively low-diversity ecological niche in young children and older adults. Importantly, relatively few bacterial genera comprise the majority of the nasopharyngeal microbiome during infancy and early childhood, with microbiome composition being influenced by breastfeeding [14, 52], mode of delivery [53, 54], acute respiratory infections [12], antibiotic treatment [12], and season [55]. Studies of older children and adolescents have primarily focused on the nasopharyngeal microbiome of individuals with asthma and other chronic respiratory diseases [56-59]. Pérez-Losada and colleagues profiled the nasopharyngeal microbiomes of 40 children and adolescents 6-17 years of age with asthma, and found that the most abundant genera were *Moraxella, Staphylococcus, Dolosigranulum, Corynebacterium, Prevotella*, and *Streptococcus* [60]. Several of these bacterial genera are reported to be highly abundant in the nasopharyngeal microbiomes of healthy adults and adults with asthma [61, 62]. To our knowledge, there are no prior studies that evaluated upper respiratory microbiome profiles from infancy through early adulthood. Our findings demonstrate that the nasopharyngeal microbiome continues to develop throughout childhood and adolescence. Future studies should investigate the biological or environmental factors that contribute to the shifts in microbiome composition that occur after early childhood.

Our study had several limitations. First, nasopharyngeal samples were collected at a single time point, and we did not have nasopharyngeal samples prior to infection in SARS-CoV-2-infected participants. Therefore, we were unable to determine if the differences in nasopharyngeal microbiome composition observed by SARS-CoV-2 infection status preceded, or were the consequence of, SARS-CoV-2 infection. Secondly, all of the SARS-CoV-2-infected study participants had relatively mild symptoms, and no study participants required antiviral treatment or hospitalization. We therefore were unable to evaluate if the microbiome features that we identified as being associated with SARS-CoV-2 respiratory symptoms are additionally associated with severe COVID-19. In addition, our use of 16S rRNA gene amplicon sequencing prevented us from evaluating other components of the upper respiratory microbiome, including viruses and fungi. Additionally, 16S rRNA gene amplicon experiments have several well-documented biases [63], although we sought to minimize these biases in our study through inclusion of all samples in a single processing run and use of appropriate negative controls. Finally, although analyses adjusted for age in evaluating associations between the nasopharyngeal microbiome and SARS-CoV-2 susceptibility and illness severity, residual confounding by unmeasured factors remains possible.

In conclusion, we found that age-associated changes in the nasopharyngeal microbiome are independently associated with SARS-CoV-2 infection and SARS-CoV-2-associated respiratory symptoms among children, adolescents, and young adults. These findings suggest that development of the nasopharyngeal microbiome during childhood and adolescence may contribute to the differences in SARS-CoV-2 susceptibility and severity observed by age. Future studies should evaluate the potential of the upper respiratory microbiome to serve as a therapeutic target for the prevention and treatment of infections caused by SARS-CoV-2 and other respiratory viruses.

## Data Availability

The sequencing dataset supporting the conclusions of this study is available in the Sequence Read Archive (PRJNA703574). The statistical files and script used for data analyses are also publicly available (https://github.com/alexmccumber/BRAVE_Kids).

## ACKNOWLEDGEMENTS

We would like to thank the Duke University School of Medicine for use of the Microbiome Core Facility, which performed the DNA extractions and library preparations for this research, and the Duke Sequencing and Genomic Technologies Core Facility, which sequenced these libraries. We offer our sincere gratitude to the children and families who participated in this research.

## DECLARATIONS

### Ethics approval and consent to participate

This study was approved by the Duke University Health System Institutional Review Board. Written informed consent was obtained from all participants or their legal guardian.

## Author contributions

JHH, AWM, SMH, MAM, JFR, CWW, and MSK contributed to the study concept and design. JNA, JR, DJL, ATR, NAT, TSP, TCG, and TND collected the data or assisted with data analysis or interpretation. JHH, AWM, and MSK drafted the manuscript and all other authors revised it critically for important intellectual content. All authors approved of the final version of the manuscript.

## SUPPLEMENTAL MATERIALS

**Supplemental Table 1**. Taxonomic classification and reference sequences for ASVs identified in nasopharyngeal samples from the study population

**Supplemental Table 2**. Differentially abundant ASVs by age in ANCOM-II analyses adjusting for SARS-CoV-2-infection status and SARS-CoV-2-associated respiratory symptoms

**Supplemental Table 3**. Prevalences and relative abundances of highly abundant bacterial genera by age

**Supplemental Table 4**. Differentially abundant ASVs by SARS-CoV-2 infection status in ANCOM-II analyses adjusting for age

**Supplemental Table 5**. Differentially abundant ASVs among SARS-CoV-2-infected individuals by the presence of respiratory symptoms in ANCOM-II analyses adjusting for age

**Supplemental Table 6**. Differentially abundant ASVs in analyses comparing SARS-CoV-2-infected individuals *with* respiratory symptoms to SARS-CoV-2-uninfected individuals in ANCOM-II analyses adjusting for age

