## Supplemental Table 3 for "Age-related changes in the upper respiratory microbiome are associated with SARS-CoV-2 susceptibility and illness severity"

**Supplemental Table 3**. Prevalences and relative abundances of highly abundant bacterial genera by age

| **Bacterial genera** | | **0-2 years** | | **3-5 years** | | **6-8 years** | | **9-11 years** | | **12-14 years** | | **15-17 years** | | **18-20 years** | |
| --- | --- | --- | --- | --- | --- | --- | --- | --- | --- | --- | --- | --- | --- | --- | --- |
| *Anaerococcus* | |  | |  | |  | |  | |  | |  | |  | |
|  | Prevalence (present/total) | 33% | (13/40) | 37% | (14/38) | 25% | (10/40) | 47% | (14/30) | 75% | (27/36) | 83% | (40/48) | 90% | (38/42) |
|  | Median (IQR) abundance | 0.00 | (0.00–0.00) | 0.00 | (0.00–0.00) | 0.00 | (0.00–0.00) | 0.00 | (0.00–0.001) | 0.01 | (0.00–0.04) | 0.01 | (0.001–0.04) | 0.02 | (0.004–0.05) |
| *Corynebacterium* | |  | |  | |  | |  | |  | |  | |  | |
|  | Prevalence (present/total) | 98% | (39/40) | 100% | (38/38) | 98% | (39/40) | 97% | (28/30) | 97% | (35/36) | 100% | (48/48) | 100% | (42/42) |
|  | Median (IQR) abundance | 0.19 | (0.06–0.36) | 0.10 | (0.02–0.29) | 0.14 | (0.03–0.34) | 0.16 | (0.03–0.41) | 0.19 | (0.10–0.47) | 0.31 | (0.09–0.44) | 0.32 | (0.20–0.66) |
| *Dolosigranulum* | |  |  |  |  |  |  |  |  |  |  |  |  |  |  |
|  | Prevalence | 98% | (39/40) | 97% | (37/38) | 98% | (39/40) | 97% | (29/30) | 83% | (30/36) | 96% | (46/48) | 93% | (39/42) |
|  | Median (IQR) abundance | 0.13 | (0.02–0.31) | 0.12 | (0.02–0.35) | 0.14 | (0.01–0.30) | 0.02 | (0.00–0.18) | 0.01 | (0.00–0.16) | 0.001 | (0.00–0.09) | 0.001 | (0.00–0.14) |
| *Fusobacterium* | |  | |  | |  | |  | |  | |  | |  | |
|  | Prevalence (present/total) | 43% | (17/40) | 55% | (21/38) | 58% | (23/40) | 63% | (19/30) | 28% | (10/36) | 44% | (21/48) | 24% | (10/42) |
|  | Median (IQR) abundance | 0.00 | (0.00–0.00) | 0.00 | (0.00–0.001) | 0.00 | (0.00–0.004) | 0.00 | (0.0–0.004) | 0.00 | (0.00–0.00) | 0.00 | (0.0–0.001) | 0.00 | (0.00–0.00) |
| *Lawsonella* | |  | |  | |  | |  | |  | |  | |  | |
|  | Prevalence (present/total) | 30% | (12/40) | 21% | (8/38) | 13% | (5/40) | 27% | (8/30) | 53% | (19/36) | 83% | (40/48) | 95% | (40/42) |
|  | Median (IQR) abundance | 0.00 | (0.00–0.00) | 0.00 | (0.00–0.00) | 0.00 | (0.00–0.00) | 0.00 | (0.00–0.00) | 0.00 | (0.00–0.01) | 0.02 | (0.002–0.08) | 0.03 | (0.01–0.06) |
| *Moraxella* | |  | |  | |  | |  | |  | |  | |  | |
|  | Prevalence (present/total) | 95% | (38/40) | 97% | (37/38) | 93% | (37/40) | 93% | (28/30) | 78% | (28/36) | 77% | (37/48) | 71% | (30/42) |
|  | Median (IQR) abundance | 0.25 | (0.00–0.53) | 0.24 | (0.02–0.66) | 0.03 | (0.00–0.20) | 0.00 | (0.00–0.05) | 0.002 | (0.00–0.13) | 0.00 | (0.0–0.004) | 0.00 | (0.00–0.01) |
| *Peptonophilus* | |  | |  | |  | |  | |  | |  | |  | |
|  | Prevalence (present/total) | 10% | (4/40) | 21% | (8/38) | 15% | (6/40) | 27% | (8/30) | 56% | (20/36) | 73% | (35/48) | 76% | (32/42) |
|  | Median (IQR) abundance | 0.00 | (0.00–0.00) | 0.00 | (0.00–0.00) | 0.00 | (0.00–0.00) | 0.00 | (0.00–0.00) | 0.00 | (0.0–0.034) | 0.009 | (0.00–0.07) | 0.01 | (0.00–0.06) |
| *Staphylococcus* | |  | |  | |  | |  | |  | |  | |  | |
|  | Prevalence (present/total) | 98% | (39/40) | 97% | (37/38) | 98% | (39/40) | 100% | (30/30) | 100% | (36/36) | 100% | (48/48) | 100% | (42/42) |
|  | Median (IQR) abundance | 0.001 | (0.00–0.003) | 0.01 | (0.00–0.06) | 0.08 | (0.002–0.42) | 0.24 | (0.04–0.69) | 0.23 | (0.06–0.52) | 0.115 | (0.04–0.30) | 0.20 | (0.08–0.42) |
| *Streptococcus* | |  | |  | |  | |  | |  | |  | |  | |
|  | Prevalence (present/total) | 98% | (39/40) | 97% | (37/38) | 93% | (37/40) | 93% | (28/30) | 86% | (31/36) | 83% | (40/48) | 76% | (32/42) |
|  | Median (IQR) abundance | 0.03 | (0.006–0.08) | 0.003 | (0.001–0.023) | 0.01 | (0.001–0.05) | 0.01 | (0.001–0.04) | 0.002 | (0.0–0.013) | 0.003 | (0.00–0.03) | 0.002 | (0.00–0.01) |

IQR–interquartile range
